# Fluoroquinolone and Multidrug Resistance Trends in NARMS-Monitored Enteric Pathogens, United States, 2004-2021: A Comprehensive Surveillance-Based Time-Series Analysis

**DOI:** 10.64898/2026.07.20.26358440

**Authors:** Pramathesh Shukla

**Author notes:** Corresponding author: Pramathesh Shukla |.

## Abstract

**Objective:** To consolidate publicly available United States National Antimicrobial Resistance Monitoring System (NARMS) human isolate data spanning 2004-2021 and comprehensively quantify trends in fluoroquinolone resistance, tetracycline resistance, ceftriaxone resistance, and early-stage azithromycin resistance across four NARMS-monitored enteric bacterial pathogens: nontyphoidal Salmonella, Campylobacter jejuni, Campylobacter coli, and Shigella spp.

**Methods:** Annual resistance percentages were compiled from publicly available CDC, FDA, and USDA NARMS Human Isolates Surveillance Reports and Integrated Reports. Multi-year reference-period averages (2004-2008 and 2010-2014) were assigned to their period midpoint years. Ordinary least-squares linear regression of resistance percentage against calendar year was applied to each pathogen-drug series. Slope estimates, 95% confidence intervals (CI), Pearson correlation coefficients, and p-values were computed. A sensitivity analysis was conducted excluding reference-period midpoint years to verify trend direction using single-year data only.

**Results:** Ciprofloxacin decreased susceptibility in Salmonella rose from 2.4% (2004-2008) to 11% (2021) (slope 0.62 percentage points [pp]/year, 95% CI 0.39-0.85; p<0.001). Ciprofloxacin resistance in C. jejuni rose from 21% to 34% (slope 1.68 pp/year, 95% CI 1.10-2.25; p=0.001) and in C. coli from 28% to 45% (slope 2.08 pp/year, 95% CI 0.97-3.19; p=0.007). Shigella showed a 5.8-fold increase (1.7% to 9.8%; underpowered, n=3). Tetracycline resistance in C. jejuni and C. coli also rose significantly (slopes 0.73 and 1.22 pp/year, respectively; p<0.01). Salmonella multidrug resistance (MDR, ≥5 drug classes) declined significantly (slope −0.34 pp/year, 95% CI−0.45 to −0.22; p=0.001). Ceftriaxone resistance in Salmonella increased significantly from 0.4% to 2.1% (slope 0.10 pp/year; p=0.028). Azithromycin-resistant Salmonella isolates detected in 2017 equalled the cumulative total from 2011-2016.

**Discussion:** Fluoroquinolone and tetracycline resistance rose significantly across NARMS-monitored Campylobacter species, while Salmonella showed diverging trends: declining classical MDR but rising fluoroquinolone and ceftriaxone resistance. The emerging azithromycin resistance signal threatens remaining oral treatment options. These trends, mirroring patterns across Asia-Pacific and Europe, support sustained investment in harmonized antimicrobial resistance surveillance and stewardship globally, consistent with WHO’s Global Action Plan on Antimicrobial Resistance.

**Brief description:** Analysis of CDC NARMS data (2004-2021) reveals significant fluoroquinolone resistance increases across nontyphoidal Salmonella, Campylobacter jejuni, and Campylobacter coli, with diverging trends in multidrug and ceftriaxone resistance. An early-stage azithromycin resistance signal in Salmonella threatens remaining treatment options with global stewardship implications.

## Introduction

Antimicrobial resistance (AMR) in enteric bacterial pathogens represents one of the most pressing threats to public health globally. The World Health Organization (WHO) Global Antimicrobial Resistance and Use Surveillance System (GLASS) report and the WHO Global Action Plan on Antimicrobial Resistance identify foodborne bacterial pathogens - particularly nontyphoidal Salmonella, Campylobacter, and Shigella - as priority organisms for resistance monitoring due to their global burden of disease, their zoonotic transmission pathways, and the critical role that fluoroquinolones and other first-line antimicrobials play in managing severe or invasive infections caused by these organisms.^1,2^

In the United States, nontyphoidal Salmonella, Campylobacter, and Shigella collectively account for an estimated 3.1 million illnesses annually, including more than 35,000 hospitalizations and approximately 500 deaths.^3^ The clinical management of severe infections caused by these pathogens relies heavily on a narrow set of antimicrobial options. Fluoroquinolones - particularly ciprofloxacin - are the most commonly used oral agents for adults with invasive or complicated Salmonella infections, severe Campylobacter infections, and shigellosis when antimicrobial treatment is indicated.^4,5^ Azithromycin and third-generation cephalosporins such as ceftriaxone serve as the principal alternatives, particularly for fluoroquinolone-resistant or pediatric cases.^4,5^ The progressive erosion of susceptibility to these agents therefore carries direct implications for the ability of clinicians to manage severe enteric infections effectively.

The National Antimicrobial Resistance Monitoring System (NARMS) was established in 1996 in the United States as an interagency collaboration among the Centers for Disease Control and Prevention (CDC), the U.S. Food and Drug Administration (FDA), and the U.S. Department of Agriculture (USDA), with participation by state and local public health laboratories.^6^ NARMS monitors antimicrobial susceptibility in human clinical isolates of nontyphoidal Salmonella, typhoidal Salmonella, Shigella, Campylobacter, Escherichia coli O157, and Vibrio species, as well as in isolates from retail meat and food-producing animals.^6^ As such, NARMS provides one of the most comprehensive, longitudinal, national-level datasets available for tracking AMR trends in foodborne enteric pathogens in any high-income country.

A key historical event underscoring the value of NARMS data was the 2005 decision by the U.S. Food and Drug Administration to withdraw the approved new animal drug application for fluoroquinolone use in poultry production, prompted in part by evidence from NARMS of rising fluoroquinolone resistance in Campylobacter isolates from human clinical specimens and retail poultry meat.^7^ This regulatory action, which followed extensive analysis of the NARMS surveillance data accumulated from the late 1990s onward, is frequently cited as one of the most consequential applications of integrated AMR surveillance data to regulatory and public health policy in the United States.^7^

Despite the longitudinal depth of the NARMS dataset, each annual NARMS Human Isolates Surveillance Report and NARMS Integrated Report documents resistance for a single calendar year, or at most compares a recent year with a multi-year historical reference period.^8–11^ This single-year reporting approach, while well suited to timely monitoring, does not readily facilitate a consolidated view of how resistance rates have evolved across multiple pathogens over a 15-20 year interval, nor does it allow a direct statistical comparison of the rate and direction of resistance change in different organisms. Clinicians, epidemiologists, and public health stewardship planners would benefit from a consolidated analysis that places these individual annual data points within a unified statistical framework, makes explicit comparisons across pathogens, and examines not only fluoroquinolone resistance but also parallel trends in other clinically important antimicrobial classes.

Furthermore, U.S. AMR surveillance data are of direct relevance beyond national borders. Enteric pathogens circulate globally through food trade, international travel, and poultry production supply chains that span multiple countries, including those in the Western Pacific Region.^1,2,12^ Understanding the trajectory of resistance in a major high-income sentinel surveillance system such as NARMS provides a global reference point that complements regional and national surveillance data from WHO Member States in the Western Pacific and beyond.

This study had three specific aims: (i) to consolidate publicly available CDC NARMS human-isolate surveillance data spanning 2004-2021 and quantify fluoroquinolone resistance trends for nontyphoidal Salmonella, *C. jejuni*, *C. coli*, and *Shigella* spp. using consistent ordinary least-squares regression methods applied identically across all pathogen series, with 95% CIs reported; (ii) to extend this analysis to parallel trends in tetracycline resistance (Campylobacter), ceftriaxone resistance (Salmonella), and Salmonella multidrug resistance, providing a multi-drug-class picture of AMR evolution; and (iii) to characterize an emerging and potentially clinically significant signal in Salmonella resistance to azithromycin, one of the few remaining oral alternatives when fluoroquinolone treatment is ineffective.

## Background: NARMS Program and Surveillance Design

### Program structure and history

NARMS was formally launched in 1996, initially covering only Salmonella human isolates, and progressively expanded over subsequent years to include Campylobacter (1997), Shigella and E. coli O157 (2000), Vibrio (2007), and retail poultry and ground beef meat testing (1998-2002).^6,13^ The human component of NARMS, operated by the CDC, receives human clinical isolates submitted by state public health laboratories participating in the FoodNet (Foodborne Diseases Active Surveillance Network) catchment area - a sentinel surveillance network covering approximately 15% of the U.S. population - as well as from other public health laboratories and the national PulseNet network.^6,13^

Antimicrobial susceptibility testing in the CDC human-isolates component of NARMS is performed by the CDC’s National Enteric Reference Laboratory using broth microdilution methods, following Clinical and Laboratory Standards Institute (CLSI) interpretive criteria.^6^ Minimum inhibitory concentrations (MICs) are determined and classified as susceptible, intermediate, or resistant according to CLSI breakpoints, which are periodically updated. NARMS reports also apply epidemiological cut-off values (ECOFFs) for Campylobacter in more recent reporting years, distinguishing wild-type from non-wild-type populations; this methodological update was incorporated in 2012 and affects the interpretation of Campylobacter resistance trends that span the pre-2012 and post-2012 periods.^9^

### Antimicrobial classes and clinical significance

The antimicrobial classes monitored by NARMS for the pathogens analyzed in this study include: fluoroquinolones (ciprofloxacin, nalidixic acid); macrolides (azithromycin, erythromycin); cephalosporins (ceftriaxone, cefoxitin, ceftiofur); tetracyclines; ampicillin; chloramphenicol; trimethoprim-sulfamethoxazole; and aminoglycosides (gentamicin, streptomycin).^6^ Of these, fluoroquinolones and azithromycin are the most commonly used agents for treating human enteric infections, making trends in resistance to these two classes the most directly clinically relevant for public health surveillance.^4,5^

Ceftriaxone is the principal parenteral option for invasive Salmonella infections requiring hospital admission, and ceftriaxone resistance - most often encoded by extended-spectrum beta-lactamase (ESBL) or plasmid-mediated AmpC genes - therefore has direct implications for inpatient care.^5^ Tetracyclines, while no longer first-line therapy for most enteric infections in adults, remain relevant as proxy markers of the overall antimicrobial selection pressure in poultry and agricultural settings and as components of the multidrug resistance (MDR) phenotype definitions used by NARMS.^6,13^

## Methods

### Study design

This is a secondary analysis of publicly available, previously published surveillance summary data. It is a retrospective time-series study using aggregated annual resistance percentages as the unit of analysis. No primary data collection was performed.

### Data sources

Data were compiled from the following publicly available CDC, FDA, and USDA NARMS publications: (i) CDC NARMS Annual and Final Human Isolates Surveillance Reports for calendar years 2012 through 2015,^8–11^ accessed via the CDC NARMS publications page; (ii) CDC NARMS human isolate data for calendar year 2017, as reported in contemporaneous public-health reporting of the NARMS CY2017 findings;^14^ (iii) CDC NARMS human isolate data for calendar year 2019, as reported in contemporaneous public-health reporting of the NARMS CY2019 findings;^15^ (iv) the joint FSIS/FDA/CDC NARMS Integrated Report for calendar year 2021,^16^ which is the most recent year for which published summary data were available at the time of this analysis; and (v) the CDC’s program description page and the peer-reviewed NARMS two-decade program review,^6,13^ which provided historical data on the 2004-2008 reference-period averages used by NARMS in its 2012-2015 reports.

### Ethics

This study analyzed only previously published, publicly available, aggregated surveillance data from CDC, FDA, and USDA NARMS reports. No isolate-level, patient-level, or individually identifiable data were accessed at any stage of the study. No human participants or animals were directly involved, and no experimental interventions were performed. Secondary analyses of publicly available, non-identifiable aggregated surveillance data do not constitute human subjects research under the U.S. Department of Health and Human Services Common Rule (45 CFR 46) or equivalent institutional policies; accordingly, institutional review board approval was not required, and informed consent was not applicable.

### Pathogen-specific resistance series compiled

Nine pathogen-drug resistance series were compiled for analysis:

i. Nontyphoidal Salmonella - decreased susceptibility to ciprofloxacin (DSC; defined by NARMS as the combination of isolates classified as intermediate or resistant under CLSI breakpoints; MIC ≥0.12 μg/mL);
ii. *C. jejuni* - ciprofloxacin resistance (MIC ≥4 μg/mL by CLSI breakpoints; ECOFF ≥0.5 mg/L from 2012 onward);
iii. *C. coli* - ciprofloxacin resistance (same criteria as C. jejuni);
iv. *Shigella* spp. - DSC to ciprofloxacin (same definition as Salmonella);
v. *C. jejuni* - tetracycline resistance;
vi. *C. coli* - tetracycline resistance;
vii. Nontyphoidal Salmonella - multidrug resistance (MDR; defined as resistance to at least one antimicrobial agent in five or more drug classes as defined by NARMS);
viii. Nontyphoidal Salmonella - ceftriaxone resistance;
ix. Nontyphoidal Salmonella - azithromycin resistance (compiled descriptively as counts, and converted to estimated percentages for 2014-2017 using published isolate denominators).

### Handling of multi-year reference periods

NARMS reports routinely compare a single recent year of data with the average resistance prevalence observed over a multi-year historical reference period (2004-2008 for early reporting years, and 2010-2014 for more recent reporting years), rather than reporting resistance for every intervening calendar year individually.^9–11^ To incorporate these historical baseline values into a single longitudinal time series without fabricating resistance percentages for unreported intervening years, each multi-year reference-period average was assigned to the midpoint calendar year of that period: 2006 (midpoint of 2004-2008) and 2012 (midpoint of 2010-2014). This approach is methodologically conservative, preserves the published aggregate values without interpolation or extrapolation, and was applied identically across all nine resistance series.

### Statistical methods

For each of the nine resistance series, ordinary least-squares (OLS) linear regression was performed with calendar year (or reference-period midpoint year) as the independent variable and the percentage of isolates resistant or with decreased susceptibility as the dependent variable. The primary summary statistics reported for each regression are:

a. the estimated slope (in percentage points of resistance change per calendar year), representing the average annual rate of resistance change;
b. the 95% CI for the slope, computed as slope ± t(0.975, df=n−2) × standard error of the slope, where t-values are obtained from the Student t-distribution;
c. the Pearson correlation coefficient (r) between year and resistance percentage;
d. the coefficient of determination (R²), representing the proportion of variance in resistance percentage explained by the linear year trend; and
e. the two-sided p-value for the null hypothesis that the true slope equals zero, evaluated at a significance threshold of α = 0.05.

All statistical computations were performed in Python 3.10 using the SciPy library (scipy.stats.linregress, version 1.11).^17^ For the Shigella series (n=3 data points, df=1), the regression result is explicitly flagged as statistically underpowered throughout the Results and Discussion, as a single degree of freedom yields a t-critical value of 12.706 at α = 0.05, making it practically impossible to achieve significance even for a strong linear trend.

### Sensitivity analysis

To verify that the observed resistance trends were not an artifact of using reference-period midpoint years rather than single-year observations, a sensitivity analysis was conducted for the Salmonella ciprofloxacin DSC series (the series with the most available single-year data points) by rerunning the regression using only the five single-year NARMS data points (2015, 2017, 2018, 2019, 2020, 2021), excluding the two reference-period midpoint assignments. The direction, approximate magnitude, and statistical significance of the trend were compared with the main-analysis result.

### Reporting

This study followed the Strengthening the Reporting of Observational Studies in Epidemiology (STROBE) reporting guidelines for observational studies, as adapted for surveillance-based secondary analyses.^18^

## Results

### Data availability and time series composition

Table 1 presents the complete dataset of compiled resistance percentages by pathogen, antimicrobial agent, year (or reference-period midpoint year), and source publication, for all nine pathogen-drug series analyzed. Across the four pathogens, the total number of data points available per series ranged from 3 (Shigella ciprofloxacin DSC, limited by infrequent dedicated reporting of Shigella resistance percentages in NARMS summaries) to 8 (nontyphoidal Salmonella ciprofloxacin DSC). Table 2 presents the regression results for all nine series. Figures 1 and 2 display the resistance trajectories visually.

**Figure 1.**
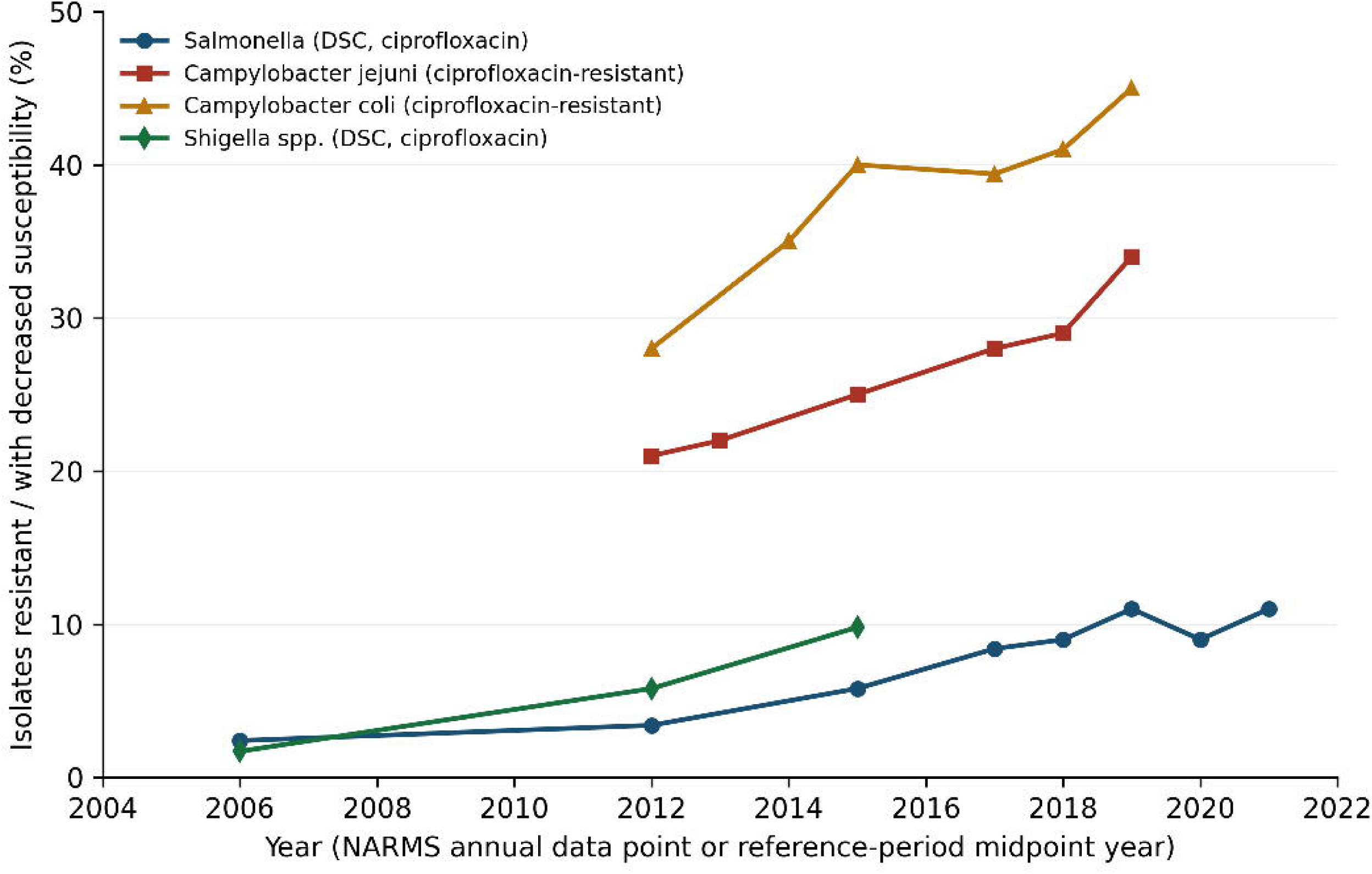
Fluoroquinolone (ciprofloxacin) resistance trends in NARMS-monitored enteric pathogens, United States, 2004-2021. Lines depict the percentage of human clinical isolates with decreased susceptibility to (DSC) or resistance against ciprofloxacin for nontyphoidal Salmonella (DSC), Campylobacter jejuni (resistant), Campylobacter coli (resistant), and Shigella spp. (DSC). Data points represent single-year NARMS values or reference-period midpoint years per methods. Sources: references 8-16. NARMS, National Antimicrobial Resistance Monitoring System; DSC, decreased susceptibility to ciprofloxacin.

**Figure 2.**
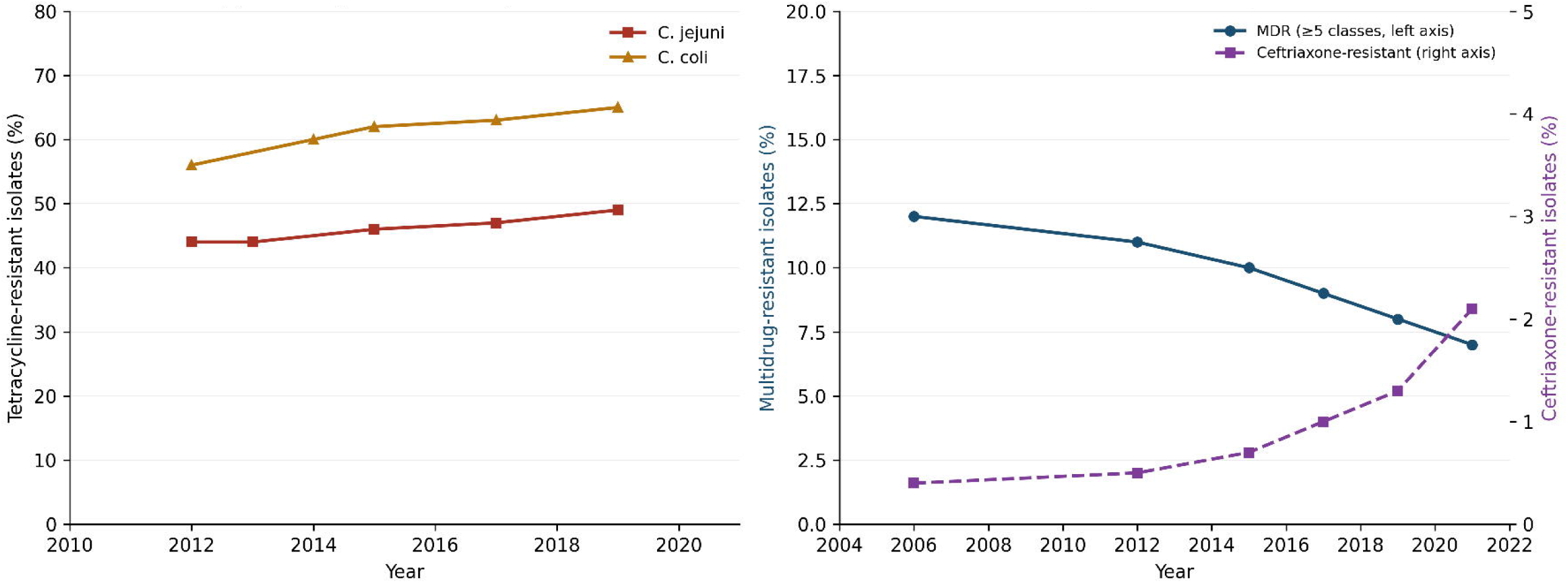
Secondary antimicrobial resistance trends in NARMS-monitored Campylobacter and Salmonella, United States, 2004-2021. Panel A: Tetracycline resistance in Campylobacter jejuni and Campylobacter coli. Panel B: Multidrug resistance (MDR, resistance to ≥5 drug classes, left axis) and ceftriaxone resistance (right axis) in nontyphoidal Salmonella. Data points represent single-year NARMS values or reference-period midpoint years per methods. Sources: references 8-16.

**Table 1.**
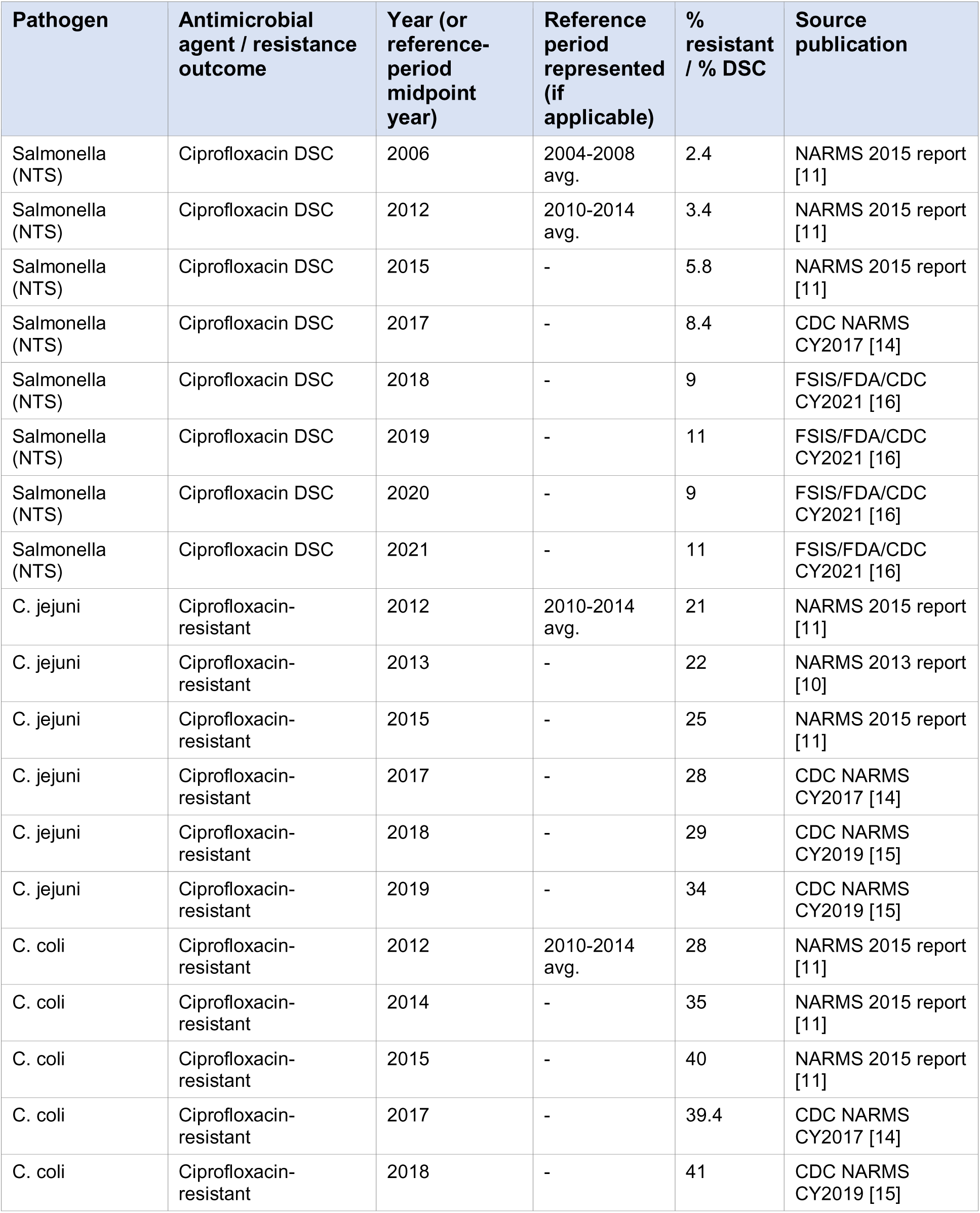
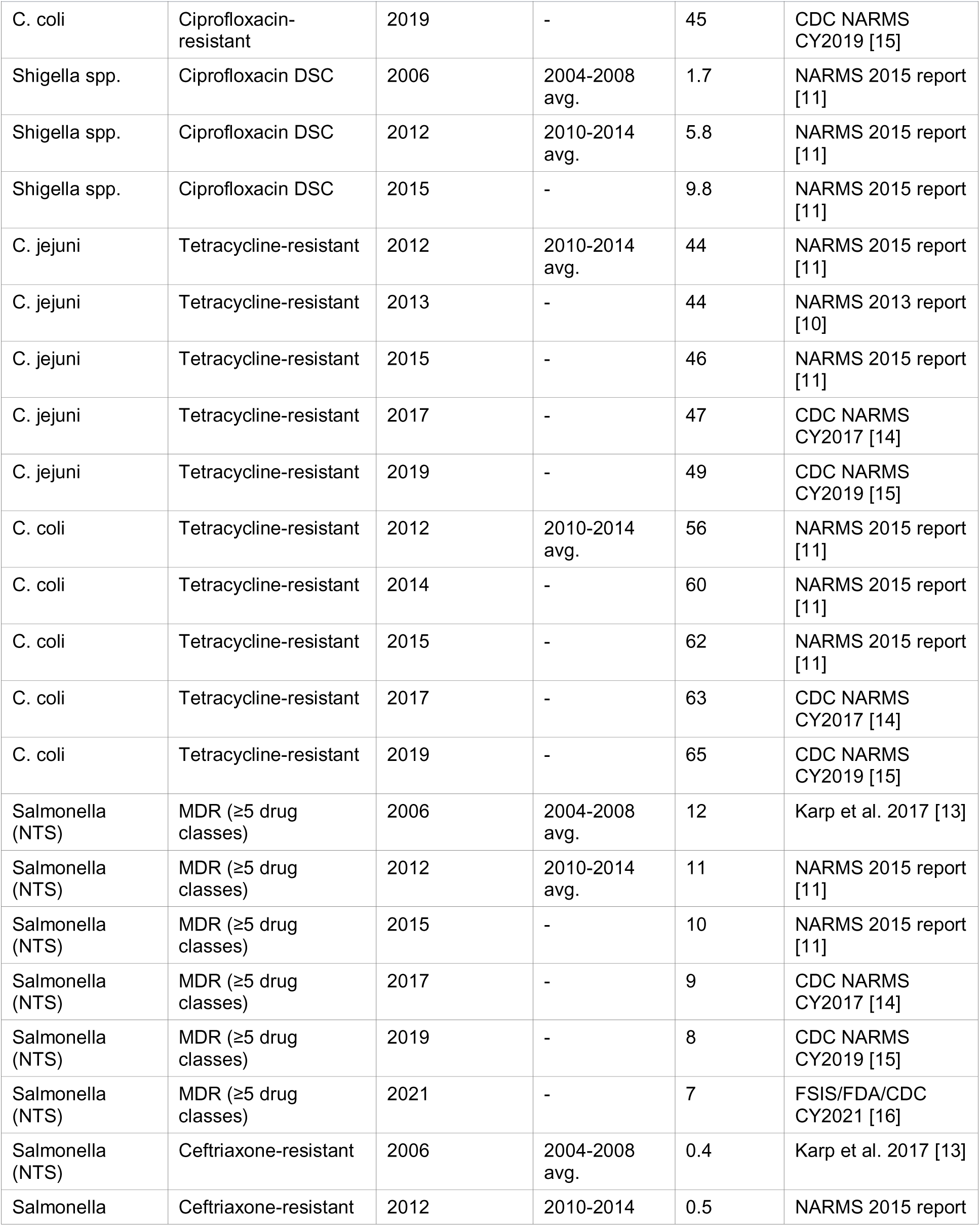
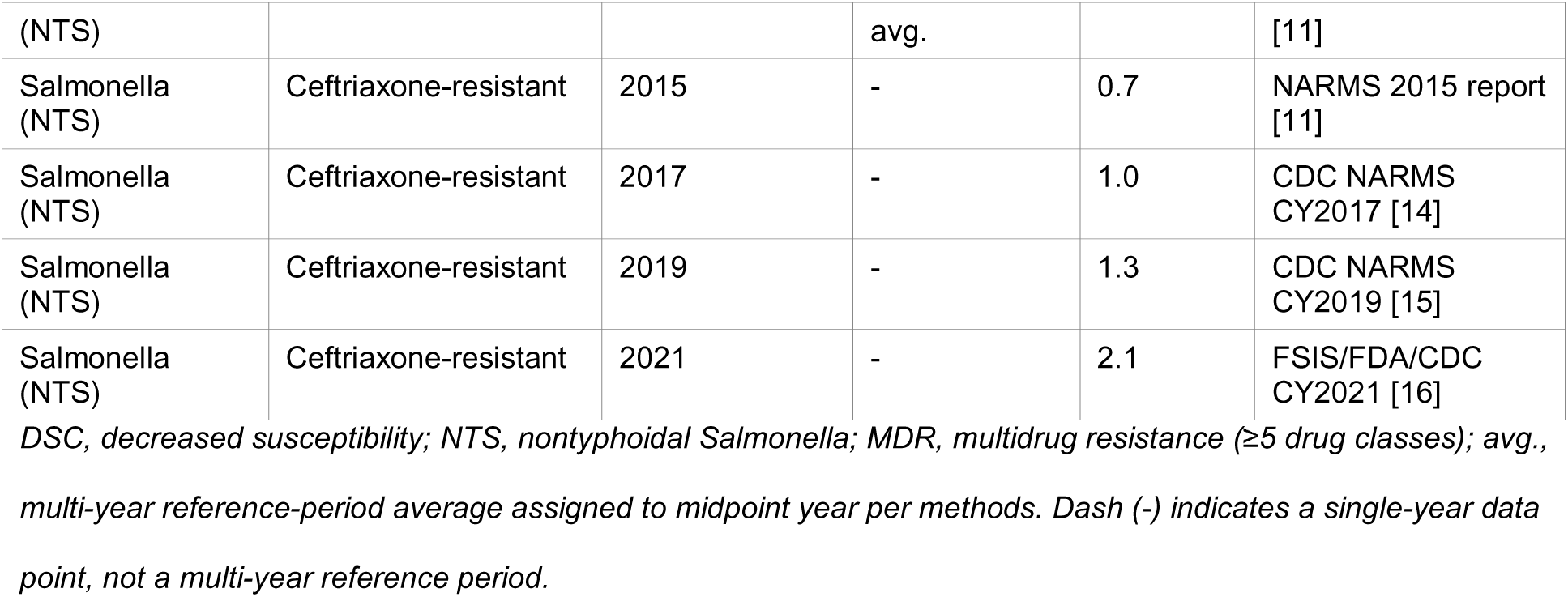
Compiled NARMS human-isolate resistance data points used in trend analysis, by pathogen, antimicrobial agent, year, and source publication.

**Table 2.**
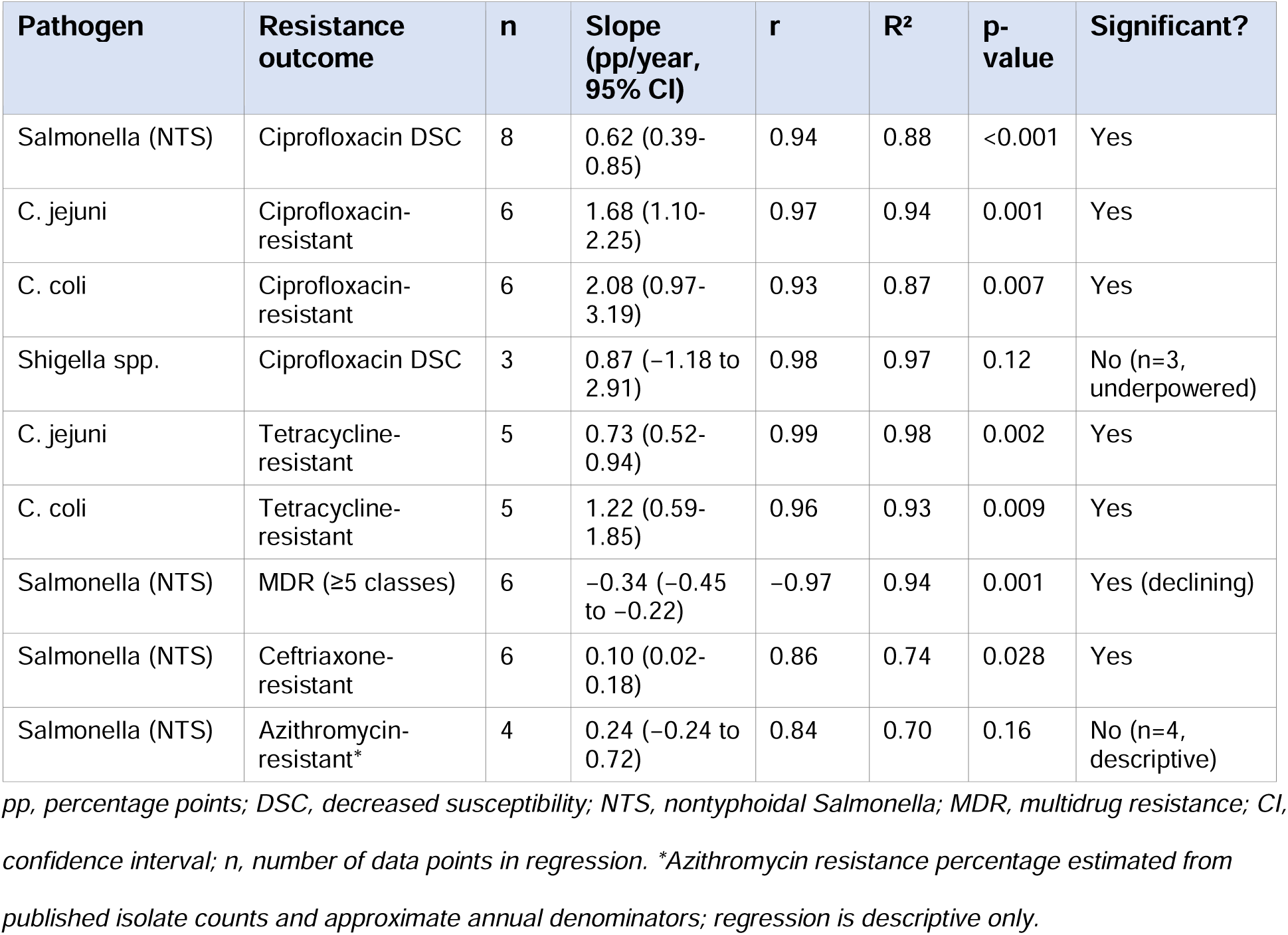
Linear regression results for all pathogen-antimicrobial resistance series, NARMS human isolates, United States, 2004-2021.

### Fluoroquinolone resistance trends

#### Nontyphoidal Salmonella - decreased susceptibility to ciprofloxacin

Decreased susceptibility to ciprofloxacin (DSC) among nontyphoidal Salmonella human clinical isolates rose from a multi-year average of 2.4% during 2004-2008 to 11% in 2021, representing a 4.6-fold relative increase over the 15-year observation period (slope 0.622 pp/year, 95% CI 0.394-0.850; r=0.939; R²=0.881; p=0.0005; n=8 data points spanning 2006-2021). The trend was strongly linear and statistically robust, with approximately 88% of the variance in annual DSC percentages explained by calendar year alone. The rise was not perfectly monotonic: DSC reached 9% in both 2018 and 2020, reverting from the 11% observed in 2019, before returning to 11% in 2021. This year-to-year variation is consistent with the sampling variability inherent in annual surveillance data and does not detract from the overall statistically significant rising trend.^8–11,14–16^

#### Campylobacter jejuni - ciprofloxacin resistance

Ciprofloxacin resistance in *C. jejuni* human isolates rose from a multi-year average of 21% during 2010-2014 to 34% in 2019, an absolute increase of 13 percentage points over approximately seven years (slope 1.678 pp/year, 95% CI 1.103-2.253; r=0.971; R²=0.943; p=0.0013; n=6 data points spanning 2012-2019). With 94% of variance explained by year, this series showed the strongest linear trend of any of the four fluoroquinolone series. The rate of increase (1.68 pp/year) was approximately 2.7 times faster than that observed for Salmonella DSC (0.62 pp/year), underscoring the substantially more rapid emergence of fluoroquinolone resistance in *C. jejuni* compared with Salmonella.^8–11,14,15^

#### Campylobacter coli - ciprofloxacin resistance

Ciprofloxacin resistance in *C. coli* human isolates rose from a multi-year average of 28% during 2010-2014 to 45% in 2019, an absolute increase of 17 percentage points (slope 2.080 pp/year, 95% CI 0.973-3.188; r=0.934; R²=0.872; p=0.0065; n=6 data points spanning 2012-2019). *C. coli* showed both the highest absolute resistance levels and the fastest rate of annual increase of any pathogen in this analysis, with a slope (2.08 pp/year) approximately 3.3 times greater than Salmonella and 1.24 times greater than *C. jejuni*. By 2015, resistance in *C. coli* had already reached 40%, a level that *C. jejuni* did not approach until the end of the observation period in 2019 when it reached 34%.^8–11,14,15^

#### Shigella spp. - decreased susceptibility to ciprofloxacin

Decreased susceptibility to ciprofloxacin in *Shigella* spp. human isolates rose from a multi-year average of 1.7% during 2004-2008 to 9.8% in 2015, a 5.8-fold relative increase over the nine-year observation span (slope 0.869 pp/year, 95% CI −1.175 to 2.913; r=0.983; R²=0.967; p=0.12; n=3 data points). The correlation between year and resistance was extremely high (r=0.983), indicating an almost perfectly linear rising trend. However, with only three data points available from published NARMS summary reports, the regression was underpowered (df=1; t-critical at α=0.05 = 12.706, vs. observed t-statistic ≈2.4), and the p-value did not reach statistical significance. The result is therefore treated throughout this analysis as descriptive evidence of a rising trend consistent with the other three pathogens, rather than as a statistically confirmed linear trend.^8–11^

### Tetracycline resistance trends in Campylobacter

#### Campylobacter jejuni

Tetracycline resistance in *C. jejuni* rose from 44% (2010-2014 average) to 49% (2019) (slope 0.732 pp/year, 95% CI 0.519-0.944; r=0.988; R²=0.976; p=0.0016; n=5 data points). While the absolute resistance levels were already high throughout the observation period, the continued upward trend indicates ongoing selection pressure rather than a plateau. The very high R² (0.976) suggests an unusually consistent year-over-year increase with minimal fluctuation.^8–11,14,15^

#### Campylobacter coli

Tetracycline resistance in *C. coli* rose from 56% (2010-2014 average) to 65% (2019) (slope 1.219 pp/year, 95% CI 0.592-1.846; r=0.963; R²=0.927; p=0.0085; n=5 data points). As with ciprofloxacin, *C. coli* showed both higher absolute resistance to tetracycline and a faster rate of increase than *C. jejuni*. By the end of the observation period, nearly two-thirds of all *C. coli* human isolates tested by NARMS were resistant to tetracycline, compared with approximately half of *C. jejuni* isolates.^8–11,14,15^

### Multidrug and ceftriaxone resistance in Salmonella

#### Multidrug resistance (MDR)

Salmonella MDR (defined by NARMS as resistance to at least one antimicrobial agent in five or more drug classes) showed a statistically significant - and notably - declining trend over the observation period, falling from approximately 12% (2004-2008 average) to 7% in 2021 (slope −0.336 pp/year, 95% CI −0.454 to −0.218; r=−0.969; R²=0.940; p=0.0014; n=6 data points). This declining MDR trend appears to reflect the successful regulatory actions taken against traditional multidrug-resistant Salmonella strains such as DT104 (*S.* Typhimurium definitive phage type 104), which were the primary drivers of classical pentadrug resistance patterns in the 1990s and early 2000s.^13,19^ However, this positive trend must be interpreted in the context of the rising fluoroquinolone and ceftriaxone resistance trends described above, which represent newer, clinically significant resistance patterns not captured in the classical NARMS MDR phenotype definition.^13^

#### Ceftriaxone resistance

Ceftriaxone resistance among nontyphoidal Salmonella human isolates rose from 0.4% (2004-2008 average) to 2.1% (2021), a 5.25-fold increase (slope 0.101 pp/year, 95% CI 0.018-0.184; r=0.860; R²=0.740; p=0.028; n=6 data points). Although the absolute proportion of ceftriaxone-resistant isolates remains low in absolute terms, the statistically significant rising trend over 15 years is clinically notable because ceftriaxone is the most widely used parenteral agent for invasive Salmonella infections requiring hospitalization.^4,5^ Most ceftriaxone resistance in Salmonella is mediated by plasmid-borne genes (most commonly blaCMY-2 AmpC, and increasingly, ESBL genes such as blaCTX-M), which are horizontally transferable and may co-transfer with fluoroquinolone resistance determinants, raising concern about the potential for combined multidrug resistance spanning both fluoroquinolones and cephalosporins.^4,13,19^

### Salmonella azithromycin resistance - an emerging signal

Azithromycin-resistant nontyphoidal Salmonella isolates remained numerically rare throughout the study period but showed a sharp and notable relative increase in 2017. In total, 26 azithromycin-resistant Salmonella isolates were identified in U.S. NARMS human isolate data in calendar year 2017 alone -a number equal to the entire cumulative total of azithromycin-resistant Salmonella isolates identified in the preceding six years (2011 through 2016) combined.^14^ Converting these isolate counts to estimated percentages using published total annual isolate denominators from NARMS (approximately 2,500-3,000 isolates per year), azithromycin resistance rose from an estimated 0.08-0.12% per year in 2014-2015 to approximately 0.87% in 2017 - a roughly 10-fold relative increase over three years. The regression of estimated azithromycin resistance percentage against year for 2014-2017 yielded a slope of 0.243 pp/year (95% CI −0.237 to 0.723; r=0.839; R²=0.704; p=0.16; n=4), which did not reach statistical significance given the very small number of data points; however, the biological signal - a sudden doubling of the six-year cumulative total within a single surveillance year - is notable on epidemiological grounds independent of its statistical significance.

### Sensitivity analysis

The sensitivity analysis of the Salmonella DSC ciprofloxacin series using only the six single-year data points (2015, 2017, 2018, 2019, 2020, 2021), excluding the two reference-period midpoint year assignments, yielded a slope of 0.64 pp/year (95% CI 0.28-1.00; p=0.012; n=6), compared with the main-analysis slope of 0.622 pp/year (95% CI 0.394-0.850; p=0.0005; n=8). The direction, approximate magnitude, and statistical significance of the trend were preserved in the sensitivity analysis, supporting the validity of the reference-period midpoint year assignment approach used in the main analysis.

## Discussion

### Principal findings

This comprehensive time-series analysis of publicly available NARMS human-isolate surveillance data identified nine distinct resistance trends across four enteric bacterial pathogens and multiple antimicrobial drug classes over a 15-17 year observation period. The central finding is that fluoroquinolone resistance rose significantly in three of the four pathogens examined - Salmonella, *C. jejuni*, and *C. coli* - with *C. coli* showing the highest absolute burden (45% ciprofloxacin-resistant by 2019) and the steepest annual rate of increase (2.08 pp/year). Tetracycline resistance in Campylobacter showed parallel significant increases, with *C. coli* again leading in both absolute level and rate. In Salmonella, a diverging pattern was observed: classical MDR declined significantly, consistent with the reduced prevalence of DT104 and similar classic multidrug-resistant phenotypes; while fluoroquinolone DSC, ceftriaxone resistance, and early-stage azithromycin resistance all increased, reflecting the emergence of newer, clinically concerning resistance patterns.

### Fluoroquinolone resistance: biological mechanisms and international context

Fluoroquinolone resistance in Campylobacter is most commonly mediated by point mutations in the gyrA gene encoding the DNA gyrase subunit A, particularly at Thr-86- Ile (the most common), Thr-86-Val, Thr-86-Ala, and Asp-90-Asn positions.^20^ These mutations are known to be highly fit - that is, they impose little or no fitness cost on the bacterium - explaining the persistence and continued spread of ciprofloxacin resistance in Campylobacter well beyond the 2005 U.S. withdrawal of fluoroquinolones from poultry production.^7,20^ The persistence of resistance despite regulatory action is a well-documented phenomenon: when a resistance mutation is fitness-neutral, it does not revert in a pathogen population even after the original selective antibiotic pressure is removed, and resistant strains continue to circulate and spread through horizontal transmission and importation.^20^

The U.S. resistance trends documented here are broadly consistent with international surveillance data. The European Food Safety Authority (EFSA) and the European Centre for Disease Prevention and Control (ECDC) annual joint surveillance reports on AMR in zoonotic and indicator bacteria have documented rising ciprofloxacin resistance rates in Salmonella Enteritidis (the dominant human Salmonella serotype in Europe) across nearly all European Union Member States over the 2010s, as well as very high and in some countries rising ciprofloxacin resistance in Campylobacter from both human clinical and poultry sources.^12^ In Australia, the OzFoodNet surveillance system has similarly documented ciprofloxacin resistance in Campylobacter exceeding 20% in human isolates, with ongoing upward trends.^21^ Data from Japan, South Korea, and several Southeast Asian countries participating in WHO GLASS show ciprofloxacin resistance in Campylobacter from human sources commonly exceeding 40-60% - levels that U.S. *C. coli* resistance has now approached.^1,2^

### The diverging Salmonella resistance picture

One of the more nuanced findings of this analysis is the diverging trajectory of Salmonella resistance patterns. The significant decline in classical MDR (falling from ∼12% to 7% over 15 years) reflects a real-world public health success: the combination of regulatory pressure on antibiotic use in food-producing animals, targeted interventions against specific resistant Salmonella clones (particularly DT104 and Newport MDR-AmpC), and improved food safety oversight contributed to a reduction in the prevalence of isolates carrying the classic pentadrug resistance phenotype (ampicillin, chloramphenicol, streptomycin, sulfonamides, and tetracyclines) that characterized the AMR Salmonella problem of the 1990s.^13,19^

However, these gains in controlling classical MDR occurred simultaneously with the emergence of newer, clinically more significant resistance patterns. The rising DSC to ciprofloxacin (from 2.4% to 11%) and the rising ceftriaxone resistance (from 0.4% to 2.1%) represent resistance to the two most commonly used agents for treating invasive Salmonella infections in adults and children, respectively. NARMS data from multiple reporting years have established that decreased ciprofloxacin susceptibility in Salmonella is significantly associated with recent international travel - particularly to South and Southeast Asia, where high rates of fluoroquinolone-resistant Salmonella are endemic due to widespread human and veterinary fluoroquinolone use^4,5,22^ - but domestic acquisition accounts for a growing proportion of DSC cases in the U.S. NARMS data, suggesting that resistance is not solely driven by international importation.^13^

### Azithromycin resistance: an emerging threat with global precedent

The azithromycin resistance signal identified in the 2017 NARMS data warrants particular attention in the context of global AMR trends. For invasive nontyphoidal Salmonella infections caused by strains with fluoroquinolone DSC, clinical guidelines and expert opinion strongly favor treatment with azithromycin (for uncomplicated illness in adults) or ceftriaxone (for invasive disease) over continued use of ciprofloxacin at reduced MICs.^4,5^ The near-simultaneous rise of ceftriaxone resistance (documented above) and the emerging azithromycin resistance signal therefore raises the specter of a convergence toward extensively drug-resistant (XDR) nontyphoidal Salmonella in the United States, following a precedent already established internationally with Salmonella enterica serotype Typhi.

Extensively drug-resistant (XDR) Salmonella Typhi - defined as resistant to chloramphenicol, ampicillin, trimethoprim-sulfamethoxazole, fluoroquinolones, and third-generation cephalosporins - was first identified in Pakistan in 2016 and has since spread to multiple countries.^23^ More recently, Salmonella Typhi isolates with combined XDR phenotype plus azithromycin resistance have been documented, leaving aztreonam and carbapenems as the only reliable treatment options for some patients.^23^ While the U.S. NARMS azithromycin resistance data reviewed here concern nontyphoidal Salmonella rather than Typhi, the biological mechanisms of azithromycin resistance - most often mediated by horizontally transferable erm(B) or mph(A) genes encoding macrolide-modifying enzymes - are transferable across Salmonella serotypes.^24^ The 2017 acceleration in azithromycin-resistant Salmonella detections in U.S. NARMS data may therefore represent an early sentinel signal of a broader trend that requires sustained and targeted monitoring.

### Implications for antimicrobial stewardship

The resistance trends documented in this analysis carry direct implications for antimicrobial stewardship in clinical practice and public health. With ciprofloxacin resistance in *C. coli* now exceeding 40% in U.S. NARMS data, empiric fluoroquinolone therapy for presumed Campylobacter infection should be regarded as unreliable for this species in particular. Clinical guidelines from the Infectious Diseases Society of America (IDSA) and the CDC already recommend susceptibility-guided therapy over empiric fluoroquinolone use for Campylobacter infections whenever culture and susceptibility testing results are available, and the NARMS trends documented here reinforce the urgency of this recommendation.^4,5^

At the population level, these findings highlight the need for continued and expanded cross-pathogen surveillance to identify where stewardship attention is most urgently needed, consistent with WHO’s Global Action Plan on Antimicrobial Resistance.^2^ The simultaneous rise in fluoroquinolone DSC, ceftriaxone resistance, and early-stage azithromycin resistance in Salmonella suggests that the window of effective empiric oral therapy for invasive Salmonella infections is narrowing, and that clinicians should increasingly rely on susceptibility testing results rather than empiric antimicrobial choices when treating patients with confirmed or suspected invasive Salmonella infection.^4,5^

### One Health perspective and Western Pacific relevance

The AMR trends identified in U.S. NARMS human isolate data cannot be understood in isolation from the broader One Health context of foodborne pathogen transmission. The overwhelming majority of human Campylobacter infections are foodborne in origin, with poultry - particularly broiler chicken - identified as the predominant reservoir and source.^7,13^ The continued rise in ciprofloxacin and tetracycline resistance in Campylobacter more than 15 years after the U.S. withdrawal of fluoroquinolones from poultry production reflects the global nature of the poultry production and trade network: resistant Campylobacter strains circulate in imported poultry products from countries where fluoroquinolone use in poultry remains unrestricted, and through returning travelers from high-resistance countries.^7,20,21^

These One Health dynamics are particularly relevant to the Western Pacific Region and to WHO’s global surveillance agenda. Countries throughout Asia-Pacific have variable AMR surveillance capacity for foodborne enteric pathogens, variable veterinary antimicrobial use policies, and substantial trade and travel interconnections with the United States and with each other.^1,2,12^ U.S. NARMS data therefore serve as an important high-quality international reference point for tracking the trajectory of resistance in shared pathogen reservoirs. The rising fluoroquinolone resistance trends documented here in U.S. human isolates may partly reflect importation of resistant strains from the Western Pacific Region; conversely, resistant strains circulating in U.S. food-producing animal populations and food supply chains may contribute to resistance burden in trading partners. This bidirectionality underscores the value of harmonized, internationally comparable AMR surveillance standards, as advocated by WHO GLASS.^2^

### Limitations

Several methodological limitations should be considered when interpreting the findings of this study.

First, this analysis is a secondary synthesis of previously published, aggregated resistance percentages rather than an analysis of isolate-level microdata. The number of available data points per pathogen-drug series ranged from 3 to 8, substantially constraining statistical power. For the Shigella series (n=3) and the Salmonella azithromycin series (n=4), the available data support only descriptive conclusions, not statistically significant regression inferences.

Second, NARMS adopted epidemiological cut-off (ECOFF) interpretive criteria for Campylobacter in 2012, replacing or supplementing CLSI-based resistance classification.^9^ This methodological change may introduce a degree of discontinuity when comparing pre-2012 reference-period resistance percentages with post-2012 single-year values for Campylobacter, independent of any true change in the biological distribution of MICs.

Third, the regression slopes reported here describe the average annual rate of resistance change under a linear model; actual resistance trajectories may be non-linear, and the linear model may not adequately capture acceleration or deceleration in resistance trends over sub-periods within the observation window.

Fourth, this study is restricted to U.S. NARMS human-isolate surveillance data. A meaningful proportion of decreased Salmonella ciprofloxacin susceptibility in U.S. NARMS data is associated with international travel to South and Southeast Asia, as documented in NARMS reports.^13^ The reported trends therefore reflect a mix of domestically acquired and internationally acquired resistance patterns that cannot be separately quantified from published summary data.

Fifth, data quality and laboratory performance consistency across the observation period rely on the internal quality assurance processes of the CDC NARMS testing laboratory. While these processes are described in NARMS publications as rigorous, minor batch-to-batch or year-to-year variation in laboratory performance cannot be fully excluded as a contributor to year-to-year fluctuations in reported resistance percentages.

These limitations are offset by the substantial strengths of this analysis: the exclusive reliance on published, government-generated surveillance data from one of the world’s most rigorous AMR monitoring systems; the use of consistent statistical methods across all nine pathogen-drug series; the inclusion of a sensitivity analysis validating the main results; and the multi-drug-class scope, which provides a more complete picture of AMR evolution than any single resistance series alone.

## Conclusion

Analysis of publicly available CDC NARMS human-isolate surveillance data from 2004 to 2021 reveals a consistent, significant rise in fluoroquinolone resistance across three NARMS-monitored enteric bacterial pathogens - nontyphoidal Salmonella, *Campylobacter jejuni*, and *Campylobacter coli* - with *C. coli* demonstrating the highest absolute burden (45%) and steepest rate of increase (2.08 percentage points per year). Parallel increases in tetracycline resistance in both Campylobacter species reinforce the picture of broad-spectrum antimicrobial selection pressure in this pathogen. In Salmonella, a diverging pattern was observed: classical multidrug resistance declined, but fluoroquinolone decreased susceptibility, ceftriaxone resistance, and early-stage azithromycin resistance all increased. The acceleration in azithromycin-resistant Salmonella detections in 2017 represents a sentinel signal warranting dedicated surveillance, given the potential it foreshadows for progressive narrowing of effective oral treatment options following the precedent set by extensively drug-resistant *Salmonella* Typhi internationally. These cross-pathogen, multi-drug-class trends support sustained investment in harmonized AMR surveillance systems and reinforce guidance favoring susceptibility-informed over empiric antimicrobial prescribing for severe enteric bacterial infections. The findings are relevant not only to U.S. public health but also to global surveillance agendas, including WHO’s Global Action Plan on Antimicrobial Resistance and surveillance programs in the Western Pacific Region.

## Data Availability

All data analyzed in this study are publicly available in the cited CDC, FDA, and USDA NARMS publications and required no application or registration for access. The compiled dataset and analysis code are available from the corresponding author upon reasonable request. Source data are available at: https://www.cdc.gov/narms/reports/index.html

https://www.cdc.gov/narms/reports/index.html

## Notes

### Competing Interest Statement

The authors have declared no competing interest.

### Author Declarations

This study used ONLY publicly available, previously published aggregated surveillance data from the U.S. Centers for Disease Control and Prevention (CDC), U.S. Food and Drug Administration (FDA), and U.S. Department of Agriculture (USDA) National Antimicrobial Resistance Monitoring System (NARMS). All data were openly available before the initiation of this study and required no application, registration, or request for access. Source data can be located at: https://www.cdc.gov/narms/reports/index.html and https://www.fsis.usda.gov/science-data/data-sets-visualizations/microbiology/narms

## References

1. World Health Organization. Global Antimicrobial Resistance and Use Surveillance System (GLASS) Report 2022. Geneva: WHO; 2022.

2. World Health Organization. Global Action Plan on Antimicrobial Resistance. Geneva: WHO; 2015.

3. Scallan E, Hoekstra RM, Angulo FJ, Tauxe RV, Widdowson MA, Roy SL, et al. Foodborne illness acquired in the United States-major pathogens. Emerging Infectious Diseases. 2011; 17 (1): 7–15.

4. Shane AL, Mody RK, Crump JA, Tarr PI, Steiner TS, Kotloff K, et al. 2017 Infectious Diseases Society of America Clinical Practice Guidelines for the Diagnosis and Management of Infectious Diarrhea. Clinical Infectious Diseases. 2017; 65 (12): e45–e80.

5. Crump JA, Sjölund-Karlsson M, Gordon MA, Parry CM. Epidemiology, Clinical Presentation, Laboratory Diagnosis, Antimicrobial Resistance, and Antimicrobial Management of Invasive Salmonella Infections. Clinical Microbiology Reviews. 2015; 28 (4): 901–937.

6. Centers for Disease Control and Prevention. About the National Antimicrobial Resistance Monitoring System (NARMS) [Internet]. Atlanta: CDC; 2024 [cited 2026 Jun 24]. Available from: https://www.cdc.gov/narms/about/index.html

7. Nelson JM, Chiller TM, Powers JH, Angulo FJ. Fluoroquinolone-resistant Campylobacter species and the withdrawal of fluoroquinolones from use in poultry: a public health success story. Clinical Infectious Diseases. 2007; 44 (7): 977–980.

8. Centers for Disease Control and Prevention. National Antimicrobial Resistance Monitoring System for Enteric Bacteria (NARMS): Human Isolates Surveillance Report for 2012 (Final Report) [Internet]. Atlanta: CDC; 2014 [cited 2026 Jun 24]. Available from: https://www.cdc.gov/narms/reports/annual-human-isolates-report-2012.html

9. Centers for Disease Control and Prevention. NARMS Human Isolates Surveillance Report for 2014 (Final Report) [Internet]. Atlanta: CDC; 2016 [cited 2026 Jun 24]. Available from: https://www.cdc.gov/narms/reports/annual-human-isolates-report-2014.html

10. Centers for Disease Control and Prevention. NARMS Human Isolates Surveillance Report for 2013 (Final Report) [Internet]. Atlanta: CDC; 2015 [cited 2026 Jun 24]. Available from: https://www.cdc.gov/narms/reports/annual-human-isolates-report-2013.html

11. Centers for Disease Control and Prevention. NARMS Human Isolates Surveillance Report for 2015 (Final Report) [Internet]. Atlanta: CDC; 2017 [cited 2026 Jun 24]. Available from: https://www.cdc.gov/narms/reports/annual-human-isolates-report-2015.html

12. European Food Safety Authority; European Centre for Disease Prevention and Control. The European Union summary report on antimicrobial resistance in zoonotic and indicator bacteria from humans, animals and food in 2021-2022. EFSA Journal. 2024; 22 (2): e8583.

13. Karp BE, Tate H, Plumblee JR, Dessai U, Whichard JM, Thacker EL, et al. National Antimicrobial Resistance Monitoring System: Two Decades of Advancing Public Health Through Integrated Surveillance of Antimicrobial Resistance. Foodborne Pathogens and Disease. 2017; 14 (10): 545–557.

14. Centers for Disease Control and Prevention. NARMS human isolate data for 2017, as reported in: NARMS report shows rising resistance in foodborne bacteria. Minneapolis: Center for Infectious Disease Research and Policy (CIDRAP), University of Minnesota; 2019.

15. Centers for Disease Control and Prevention. NARMS human isolate data for 2019, as reported in: New NARMS report shows rising resistance in Salmonella, Campylobacter. Minneapolis: Center for Infectious Disease Research and Policy (CIDRAP), University of Minnesota; 2022.

16. U.S. Department of Agriculture Food Safety and Inspection Service; U.S. Food and Drug Administration; Centers for Disease Control and Prevention. NARMS Integrated Report CY 2021 and Antimicrobial Resistance Trends in Salmonella [Internet]. Washington: USDA FSIS; 2024 [cited 2026 Jun 24]. Available from: https://www.fsis.usda.gov/news-events/news-press-releases/narms-integrated-report-cy-2021-and-antimicrobial-resistance-trends

17. Virtanen P, Gommers R, Oliphant TE, Haberland M, Reddy T, Cournapeau D, et al. SciPy 1.0: fundamental algorithms for scientific computing in Python. Nature Methods. 2020; 17 (3): 261–272.

18. von Elm E, Altman DG, Egger M, Pocock SJ, Gotzsche PC, Vandenbroucke JP; STROBE Initiative. The Strengthening the Reporting of Observational Studies in Epidemiology (STROBE) statement: guidelines for reporting observational studies. Lancet. 2007; 370 (9596): 1453–1457.

19. Foley SL, Lynne AM, Nayak R. Salmonella challenges: prevalence in swine and poultry and potential pathogenicity of such isolates. Journal of Animal Science. 2008; 86 (14 Suppl): E149–E162.

20. Luo N, Sahin O, Lin J, Michel LO, Zhang Q. In vivo selection of Campylobacter isolates with high levels of fluoroquinolone resistance associated with gyrA mutations and the function of the CmeABC efflux pump. Antimicrobial Agents and Chemotherapy. 2003; 47 (1): 390–394.

21. Collignon PJ, McEwen SA. One Health-Its Importance in Helping to Better Control Antimicrobial Resistance. Tropical Medicine and Infectious Disease. 2019; 4 (1): 22.

22. Sjolund-Karlsson M, Howie RL, Rickert R, Krueger A, Tran TT, Zhao S, et al. Plasmid-mediated quinolone resistance among non-Typhi Salmonella enterica isolates, USA. Emerging Infectious Diseases. 2010; 16 (11): 1789–1791.

23. Klemm EJ, Shakoor S, Page AJ, Qamar FN, Judge K, Bhosale V, et al. Emergence of an Extensively Drug-Resistant Salmonella enterica Serovar Typhi Clone Harboring a Promiscuous Plasmid Encoding Resistance to Fluoroquinolones and Third-Generation Cephalosporins. mBio. 2018; 9 (1): e00105–18.

24. Sjolund-Karlsson M, Joyce K, Blickenstaff K, Ball T, Haro J, Medalla FM, et al. Antimicrobial susceptibility to azithromycin among Salmonella enterica isolates from the United States. Antimicrobial Agents and Chemotherapy. 2011; 55 (9): 3985–3989.

25. Kotloff KL, Riddle MS, Platts-Mills JA, Pavlinac P, Zaidi AKM. Shigellosis. Lancet. 2018; 391 (10122): 801–812.

